# Development and evaluation of a scalable alternative to chart review for phenotype case adjudication using standardized structured data from electronic health records

**DOI:** 10.1101/2022.12.27.22283944

**Authors:** Anna Ostropolets, George Hripcsak, Syed A Husain, Lauren Richter, Matthew Spotnitz, Ahmed Elhussein, Patrick B Ryan

## Abstract

**Objective:** Chart review as the current gold standard for phenotype evaluation cannot support observational research at scale. It is expensive, time-consuming, and variable. We aimed to evaluate the ability of structured data to support efficient patient status ascertainment and develop a standardized and scalable alternative to chart review.

**Methods:** We developed Knowledge-Enhanced Electronic Patient Profile Review system (KEEPER) that extracts a patient’s structured data elements relevant to a given phenotype and presents them in a standardized fashion that follows clinical reasoning principles. We evaluated its performance compared to manual chart review for four conditions (diabetes type I, acute appendicitis, end stage renal disease and chronic obstructive lung disease) using randomized two-period, two-sequence crossover design. Inter-method agreement, inter-rater agreement, accuracy, and review duration were measured.

**Results:** Ascertaining patient status with KEEPER was twice as fast compared to manual chart review. 88.1% of the patients were classified concordantly using full chart and KEEPER, but agreement varied depending on the condition. Pairs of clinicians agreed in classification of patient status in 91.2% of the cases when using KEEPER compared to 76.3% when using full chart. Patient classification aligned with the gold standard in 88.1% and 86.9% of the cases respectively.

**Conclusion:** This proof-of-concept study demonstrated that structured data can be used for efficient patient ascertainment if are limited to only relevant subset and organized according to the clinical reasoning principles. A system that implements these principles can achieve similar accuracy and higher inter-rater reliability compared to chart review at a fraction of time.

## BACKGROUND

Phenotyping algorithms or executable algorithms for identifying patients of interest in observational data are the backbone of observational research [1]. The validity of inference from observational data highly depends on their accuracy, which is commonly evaluated using manual chart review. This process is time- and labor-consuming, requires heavy clinician involvement, and is variable. Due to these limitations, phenotypes are commonly borrowed from the literature based on their previously demonstrated performance [3]. Nevertheless, good performance on one data source does not guarantee portability to another [4,5]. A need to evaluate new phenotypes and re-evaluate previous is a significant obstacle in large-scale observational research and remains the bottleneck in both data-driven and rule-based phenotyping [2].

If evaluation is performed, the researchers typically review a small subset of patients identified by the algorithm, classify each patient as true positive or false positive and estimate positive predictive value omitting sensitivity and specificity [6,7]. Incomplete validation can lead to the choice of suboptimal phenotypes. A smaller number of studies that focus on identifying the best-performing algorithm for future clinical studies examine a larger sample size but take months and require significant resources [8–11], which is not scalable for more than one condition at a time.

As only a small sample is typically reviewed, validation results can suffer from selection bias. Previous research showed that the records of those patients who consented to supply their information differed from those who did not [12]. Condition-specific phenotype-guided chart sampling strategies were proposed to account for bias during sampling. Simulation experiments showed similar statistical power with fewer charts reviewed [13,14], but the methods need to be developed further to demonstrate scalability and generalizability.

Another challenge that undermines the validity of manual chart review is variability in review strategies that are influenced by differences in training, high volume of information in health records and chart sparsity [15–17]. If patients are observed in the system regularly, the information volume grows quickly with conflicting information found in different places in the chart [18]. On the other hand, most of the content in charts is highly redundant and useful information can be buried under duplicated notes [19]. With the advances in data extraction and mining [20–22], a growing body of literature uses various natural language processing techniques to extract diagnostic information [23–28]. While these models show high flexibility and adaptability, they tend to be disease-specific, which limits their scalability.

Chart review often requires acquiring additional access to identified unstructured data, which may not be feasible for some researchers or institutions. It is not possible on the data sources with no charts such as administrative claims. While the latter offer more comprehensive patient capture as insurance tracks patients across all institutions, the inference from claims can be perceived as inferior to electronic health record (EHR) because traditional phenotype validation is not performed. One potential solution is using linked EHR-claims or registry-claims data sources where the former can act as a gold standard [29]. This type of validation is only available in a rather small number of institutions that have linked data sources. Alternatively, predictive models have been proposed to generate a probabilistic gold standard, use it to assign the probability of being a case to each patient identified by the algorithm and derive performance metrics [30]. While very promising, such approaches may lack interpretability and transparency, while reviewing charts provide an important ability to construct narratives about patients [31].

We propose that the true patient state is latent in structured data and the latter can be used to effectively ascertain patient status for phenotype evaluation. We hypothesize that three principles are crucial in this process: (a) organization of the data in the way that mimics a typical clinical diagnostic process, (b) presentation of only relevant information as opposed to the whole volume of patient structured data and (c) standardization of information extraction and representation.

We use these principles to design and evaluate a scalable and interpretable chart review alternative: Knowledge-Enhanced Electronic Profile Review system (KEEPER).

## METHODS

We will describe KEEPER, its principles, application to four conditions of interest and evaluation.

### Data source

In this study, we use Columbia University Irving Medical (CUIMC) EHRs translated to Observational Health Data Sciences and Informatics (OHDSI) Observational Medical Outcomes Partnership (OMOP) Common Data Model (CDM) [32].

CUIMC database comprises electronic health records on more than 6 million patients and includes inpatient and outpatient care. The database currently holds information about the person (demographics), visits (inpatient and outpatient), conditions (billing diagnoses and problem lists), drugs (outpatient prescriptions and inpatient orders and administrations), devices, measurements (laboratory tests and vital signs), and other observations (symptoms). The data sources include current and previous electronic health record, administrative and ancillary systems.

CUIMC OMOP CDM instance contain the structured data about patient demographics, visits, conditions, laboratory tests and measurements, diagnostic and treatment procedures as well as inpatient drug administrations and outpatient prescriptions. Some of the elements not captured in our OMOP structured data include results of imaging studies, bacterial culture tests or content of free-text notes.

### Principles

#### 1. Adherence to clinical reasoning

KEEPER applies general principles and steps of diagnostic clinical reasoning to patient structured data within the context of the phenotype being evaluated. We look at the outcome for which the phenotype was developed as a diagnosis clinicians are evaluating in a patient. We use the following elements of diagnostic reasoning to organize the extracted structured data: clinical presentation (complaints, signs, symptoms and physical examination), history (disease history, co-morbidities, risk factors and exposures), preliminary diagnosis, subsequent diagnostic procedures, diagnoses, treatment, follow-up care and complications.

#### 2. Standardization

Both data extraction and representation are standardized across data sources and conditions. Standardized extraction is supported by a common data model (in our case, OMOP CDM) and standardized representation is based on the conceptual elements described above. As the steps of clinical reasoning are universal for any condition [33], the structure of data representation is unified and, as a result, disease-agnostic.

#### 3. Dimensionality reduction

As the patient data are reviewed for the purpose of phenotype evaluation, we only extract the information that is clinically relevant to a given phenotype. We hypothesize that the structured data provides sufficient information to ascertain patient status even despite the data loss observed when when using only structured data [34].

### Conceptual elements and data elements

KEEPER is built around the conceptual elements representing the typical steps clinicians follow when diagnosing a patient, which are contextualized around a disorder of interest (Table 1).

**Table 1.** Conceptual elements and data representation in KEEPER.

| Conceptual element | Conceptual element in the context of the disease of interest | Data element |
| --- | --- | --- |
| Clinical presentation | Presence of relevant [known to be associated with the outcome] symptoms on the encounter (index date, day 0) and absence of competing symptoms | Condition codes [day 0] |
| Clinical plausibility | Appropriate demographics | Age, gender, race and ethnicity<br>[day 0] |
|  | Presence of relevant symptoms, diagnoses or treatment prior to the index date, especially recurring | Condition, drug and observation codes<br>[before day 0] |
|  | Presence of relevant co-morbidities and (or) pre-disposing risk factors | Condition and observation codes<br>[before day 0] |
|  | Absence of competing diagnoses after the index date, especially if followed by treatment | Condition, procedure, measurement and drug codes<br>[after day 0] |
| Diagnostic procedures | Presence of diagnostic procedures, laboratory tests, clinical consults with other specialties, transfer to specific care sites around the index date | Procedure codes<br>[before and after day 0] |
|  |  | Measurement codes and values<br>[before and after day 0] |
|  |  | Provider and location<br>[before and after day 0] |
| Treatment procedures and medications | Presence of relevant instrumental and surgical procedures performed on or after the index date | Procedure codes<br>[after day 0] |
|  | Presence of relevant medications prescribed or administered on or after the index date | Drug codes<br>[after day 0] |
| Follow-up care and complications | Presence of relevant follow-up visits | Provider and location<br>[after day 0] |
|  | Presence of relevant complications after the index date | Condition codes<br>[after day 0] |

The first element is clinical presentation, which consists of patient symptoms, signs, and complaints on the day they seek care (day 0 or index date). In clinical practice, physician (or healthcare team) collects current complaints, past personal and family history, assesses vital signs, performs physical examination and, based on the totality of information, makes a preliminary diagnosis.

For example, in the context of acute appendicitis phenotype, Patient X with suspected acute appendicitis (in textbook scenario) presents to the emergency room complaining of epigastric pain migrating to right lower quadrant, nausea and vomiting. Physical exam reveals fever, localized tenderness in the right lower quadrant and positive Rovsing’s sign [35].

On the data level, it translates into condition codes for corresponding signs and symptoms (such as ICD-10(CM) R11.0 ‘Nausea’), measurement codes for vital signs (such as high body temperature) or condition codes for acute appendicitis. Observing these data elements increases one’s confidence in the diagnosis and observing symptoms typical for other conditions (such as intermittent severe pain that waxes and wanes in renal colic) or competing diagnoses (diverticulitis or renal colic) decreases one’s confidence.

Next, we assess clinical plausibility, which includes specific demographics if a condition is known to be prevalent in a given group, history of disease and pre-disposing factors. Within the context of acute appendicitis phenotype, Patient X is more likely to be young [36] and less likely to have prior recurrent abdominal symptoms or have been diagnosed with Crohn’s disease or endometriosis. If a condition of interest was chronic or had known risk factors, we would expect to observe prior episodes of care or relevant comorbidities. On contrary, observing a differential diagnosis recorded after the encounter (such as Crohn’s disease), especially followed by the subsequent treatment would decrease our confidence in the diagnosis.

The next conceptual element encompasses diagnostic procedures and laboratory tests. In our clinical scenario, Patient X is sent for blood work and diagnostic imaging of the abdomen (ultrasound or computer tomography). Diagnostic findings include leukocytosis and radiographic signs of appendicitis (enlarged appendix with wall thickening or perforated appendicitis). From the data perspective, observing these diagnostic procedures along with corresponding laboratory values would increase our confidence in the diagnosis.

Treatment procedures and medications are approached in the same way. Subsequent treatment can include a short course of antibiotics (e.g., piperacillin-tazobactam or cephalosporins in combination with metronidazole), appendectomy within a day or interval appendectomy. In our scenario, Patient X undergoes laparoscopic appendectomy and pathologic examination of the appendix reveals gangrenous appendicitis. Since the final pathologic diagnosis is consistent with acute appendicitis, the clinical case can be concluded. As pathology and operative reports are oftentimes not available in the structured data, observing relevant treatment and complications of appendicitis along with absence of competing treatment (such as colectomy or gastrotomy) would conclude the case in the structured data.

Table 2 shows the examples of KEEPER for three patients with suspected acute appendicitis. The records do not reflect real patient data but are constructed based on the data from the cases we ascertained. The first patient in Table 2 (green) is 46 year old male, admitted with abdominal pain, enlarged liver and leukocytosis. Clinical presentation is consistent with acute appendicitis or umbilical hernia, so the patient is referred to computer tomography of abdomen and is treated with a short course of antibiotics. Subsequently, the patient is diagnosed with acute gangrenous appendicitis and undergoes appendectomy. Presence of relevant symptoms, diagnostic and treatment procedures and absence of competing diagnoses after the index date is highly suggestive of acute appendicitis.

**Table 2.** Examples of KEEPER for three patients with suspected acute appendicitis: likely a case (green), likely a control (red) and ambiguous (blue).

| Demographics and details about the visit | Presentation | Prior conditions, symptoms and treatment | Diagnostic procedures | Laboratory tests | Competing diagnoses | Treatment procedures and medications | Complications |
| --- | --- | --- | --- | --- | --- | --- | --- |
| Male, 46 yo;<br>Visit:<br>emergency room followed by hospitalization (3 days) | Abdominal pain; Acute appendicitis; Large liver; Umbilical hernia without obstruction AND without gangrene | Abdominal pain (day - 71);<br>Abdominal pain (day -1); | Computed tomography, abdomen and pelvis; with contrast material(s (day 0); | Leukocytes (abnormal, high, day 1); Neutrophils (normal, day 1); Neutrophils/100 leukocytes (abnormal, high, day 1) |  | Appendectomy (day 25); metronidazole (3 days) | Acute gangrenous appendicitis (day 25);<br>Acquired absence of organ (day 25) |
| Female, 17 yo;<br>Visit:<br>Hospitalization (7 days) | Abdominal pain; Appendicitis; Diverticulitis of colon; Fever; | Diverticulitis of colon (day -182); | Computed tomography, abdomen; with contrast material(s); Computed tomography, pelvis; with contrast material(s) (day 5); | Leukocytes (abnormal, high, day 0/1/2/5); Leukocytes (normal, day 3/4/6/7); Neutrophils/100 leukocytes (normal, day 0/6); Neutrophils/100 leukocytes (abnormal, high, day 1-5) | Diverticulitis of colon (day 20); | piperacillin and tazobactam (5 days); |  |
| Male, 70 yo;<br>Visit:<br>emergency room followed by hospitalization (2 days) | Acute appendicitis; Barrett's esophagus; Esophagitis; Gastrointestinal hemorrhage; Hematemesis; | Abdominal pain (day - 816);<br>Esophagitis (day -180); | Esophagogastro-duodenoscopy, flexible, transoral; diagnostic, including collection of specimen(s) by brushing or washing, when performed (day 0); | Leukocytes (abnormal, high, day -1 and 0); Leukocytes (normal, day 1); Neutrophils (normal, day -1); Neutrophils/100 leukocytes (normal, day - 1) | Diaphragmatic hernia; Barrett's esophagus; Hematemesis; Eosinophilic esophagitis; Gastrointestinal hemorrhage | pantoprazole (62 days); famotidine (2 days); ondansetron (1 days) |  |

On contrary, the last patient in Table 2 (in red) is likely a control. 70-year-old man presented to the emergency department with symptoms suggestive of an acute abdominal problem (acute appendicitis, Barrett’s esophagus and esophagitis). Given presence of hematemesis (a serious potentially life-threatening acute event with clear unambiguous presentation), we can suspect that hematemesis was the main complaint and acute appendicitis was a rule-out diagnosis.

Subsequent diagnostic procedures (presence of esophagogastroduodenoscopy for hematemesis and absence of computer tomography for appendicitis) and treatment (acid-reducing drugs) likely confirm that this patient did not have acute appendicitis.

The other patient has the elements suggestive of appendicitis (laboratory findings and appropriate treatments) but also has the elements indicative of another condition (history of diverticulitis and subsequent diagnosis of diverticulitis), so the choice regarding the status of such patient is left to the reviewer’s discretion.

Examples of conceptual elements for other conditions are provided in Supplementary Table 1.

### Experiment

As a proof-of-concept study, we implemented KEEPER for four conditions and conducted a randomized standardized experiment comparing the performance of knowledge-enhanced patient profiles and manual chart review. We selected conditions that represent chronic and acute conditions, rare and prevalent, those that are usually managed in inpatient and outpatient settings: acute appendicitis, diabetes mellitus type I (DMI), chronic obstructive pulmonary disorder (COPD), and end stage renal disease (ESRD).

#### Data extraction and gold standard

For each disease, we used eMERGE PheKB algorithms that were developed and validated on CUIMC data [37– 41]. We executed them in CUIMC EHR, selected a random subset of 20 patients for each condition and extracted relevant data elements in a semi-automated fashion.

Demographic characteristics and recorded symptoms, signs, and diagnoses on day 0 were extracted from OMOP CDM *person* and *condition_occurrence* tables without any modification. Relevant co-morbidities, disease history (recorded any time before the index date), differential diagnoses and complications (any time after the index date) were extracted from *condition_occurrence* table, where selection was guided by the SNOMED-CT hierarchy and refined iteratively based on the distribution of the concepts in CUIMC EHR data. For example, for acute appendicitis we extracted all descendants of SNOMED-CT ‘Disorder of abdomen’, ‘Disorder of pelvis’ and ‘Disorder of the genitourinary system’. Risk factors such as smoking for COPD were extracted from *observation* table. Relevant drugs (recorded any time on or after the index date) were extracted using the joint ATC-RxNorm hierarchy using grouping terms in ATC (for example, all descendants of ATC ‘Antiinfectives for systemic use’ and ‘Alimentary tract and metabolism’ for acute appendicitis) and presented at the ingredient level with days supply.

Procedures and measurements (laboratory tests and vitals recorded before, on and after the index date) were defined in groups based on clinical expertise. The codes can be found on GitHub [42].

The datasets for four conditions were then assembled similarly to Table 2 and saved as flat files. Data extraction was performed uniformly for all patients prior to their ascertainment.

Chart review was performed on full patient medical records by two authors (AO and GH) separately, labels for each patient were compared and iterative chart review continued until all disagreements were resolved.

#### Patient review

The experiment was conducted by four independent clinicians in two rounds (Figure 1). Two clinicians reviewed the patients with suspected acute appendicitis and patients with suspected DM1 and the other two – patients with suspected COPD and ESRD.

**Figure 1.**
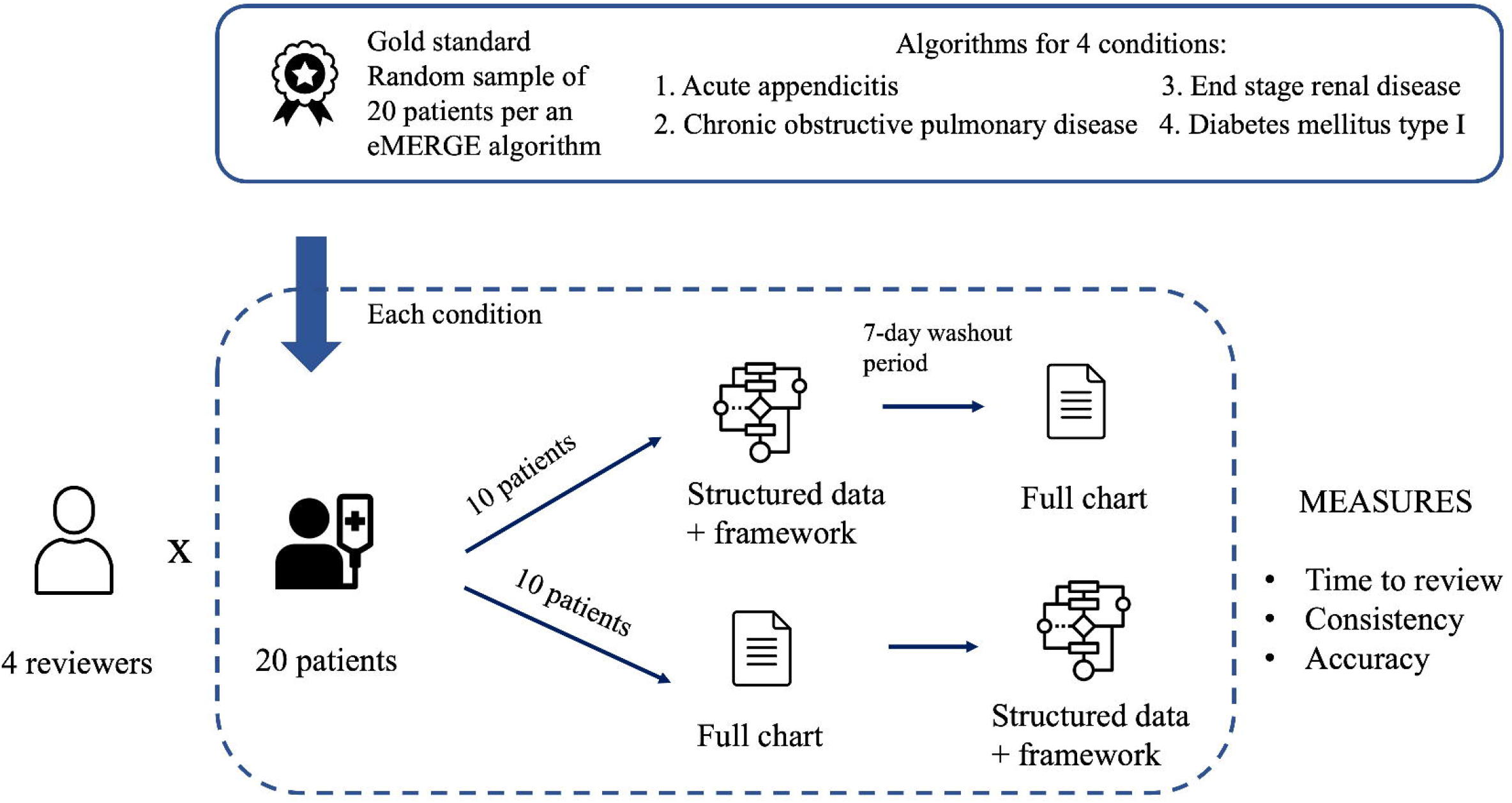
Overview of the proof-of-concept experimental design for comparing KEEPER and chart review for phenotype evaluation.

We followed two-period, two-sequence crossover design, where two-period refers to two rounds and two-sequence refers to the order of studied methods [43]. For each condition, we randomly split the patients into two groups of ten, so that during the first round a clinician reviewed the profiles of patients 1-10 and charts of patients 11-20 and during the second round – profiles of patients 11-20 and charts of patients 1-10. There was a minimum of a 7-day wash-out period between rounds. Patients were assigned different identifiers to prevent carryover effect.

Each patient was classified based on the presence of the disease of interest anytime in the patient’s history and the date the disorder was first observed in clinical settings was compared to the phenotype index date.

#### Metrics

First, we calculated the proportion of patients classified concordantly by chart review and patient profile review (inter-method agreement). We used Cohen’s kappa (chance-corrected agreement) to measure the agreement between patient profiles and charts for each condition as well as the overall agreement.

Second, we measured inter-rater agreement between two clinicians to assess if consistency of patient ascertainment among reviewers is improved by using standardized patient profiles. As we used fully crossed design with the goal of estimating reliability of the ratings from multiple clinicians, Fleiss’s kappa was chosen as the metric for the overall agreement and Cohen’s kappa for pairwise comparison [44]. The Cochran-Mantel-Haenszel test was used to compare methods across different conditions followed by Fisher exact test for pairwise comparisons [45].

Third, we compared the accuracy of ascertainment against the gold standard when using full charts and KEEPER, where the accuracy was calculated as the proportion of the labels that agree with the gold standard. Proportions were compared using the Cochran-Mantel-Haenszel test. Additionally, we compared the time to review patient profiles and full charts using the Student’s t-test and performed qualitative analysis of the discrepancies in case ascertainment.

## RESULTS

### Agreement and accuracy

We observed substantial agreement between the results of chart review and patient profile review (Table 3). Overall, 88.1% of the patients were classified similarly using full chart and KEEPER, which corresponded to Cohen’s kappa of 0.71 (95% confidence interval [CI] 0.59 – 0.83).

**Table 3.** Comparison of chart review and KEEPER: inter-method agreement, inter-rater agreement, and accuracy.

|  | Inter-method agreement |  | Inter-rater agreement |  |  |  | Accuracy |  |
| --- | --- | --- | --- | --- | --- | --- | --- | --- |
|  | Cases, n (%) | Kappa (95% CI) | Chart, n (%) | Kappa (95% CI) | KEEPE R, n (%) | Kappa (95% CI) | Chart, n (%) | KEEPE R, n (%) |
| DMI | 32 (80.0) | 0.58 (0.34-0.82) | 14 (70.0) | 0.40 (<0.1-0.78) | 18 (90.0) | 0.77 (0.47-1.00) | 34 (85.0) | 35 (87.5) |
| Acute appendicitis | 38 (95.0) | 0.87 (0.69 - 1.00) | 19 (95.0) | 0.86 (0.56 – 1.00) | 19 (95.0) | 0.88 (0.64-1.00) | 39 (97.5) | 39 (97.5) |
| COPD | 34 (85.0) | 0.67 (0.44-0.90) | 16 (80.0) | 0.60 (0.28-0.92) | 20 (100.0)<sup>+</sup> | 1.00 (1.00-1.00) | 34 (85.0) | 32 (80.0) |
| ESRD | 37 (92.5) | 0.78 (0.54-1.00) | 12 (60.0) | -0.1 (-0.3-0.1) | 15 (75.0) | 0.34 (-0.01-0.72) | 32 (80.0) | 35 (87.5) |
| Overall | 141 (88.1) | 0.71 (0.59-0.83) | 61 (76.3) | 0.45 (0.23-0.67)* | 73 (91.2)<sup>+</sup> | 0.74 (0.52-0.96)* | 139 (86.9) | 141 (88.1) |

\* indicates Fleiss's kappa to account for two pairs of reviewers; others kappas are Cohen's kappa
<sup>+</sup> indicates significant difference between two methods based on Cochran-Mantel-Haenszel test and Fisher exact test (alpha = 0.05)
Kappa ≤ 0 indicates no agreement; 0.01–0.20 - none to slight; 0.21–0.40 – fair; 0.41– 0.60 – moderate; 0.61– 0.80 – substantial; and 0.81–1.00 – almost perfect agreement

For all conditions, KEEPER provided sufficient information to arrive at the same conclusions regarding patient status as with using full charts in 80% of the cases. Agreement varied across conditions with lowest agreement between two methods for diabetes mellitus type I (moderate agreement) and highest agreement for acute appendicitis (almost perfect agreement).

When comparing inter-rater agreement (agreement in patient ascertainment between two reviewers), we observed that KEEPER enabled more consistent review. Clinicians arrived at the same conclusions regarding the patients’ status in 91.2% of the cases when using KEEPER compared to 76.3% when using full charts. This trend was observed for most of the conditions (diabetes mellitus type I, end stage renal disorder and chronic obstructive pulmonary disorder). In acute appendicitis, the reviewers achieved similar inter-rater agreement when using charts and using KEEPER.

Clinicians achieved similar accuracy of patient classification when using KEEPER compared to charts. Overall, in 88.1% and 86.9% of cases, respectively, patient classification aligned with the gold standard. In all conditions, accuracy of KEEPER was at least 80% and in three out of four conditions the accuracy was higher (albeit non-significant) or similar to the accuracy of full chart review.

### Efficiency

The use of KEEPER reduced the time needed for review in more than half in both rounds. On average, chart review for 20 patients took 67 minutes (SD = 43) and patient profile review took 30 minutes (SD = 14, p-value 0.04).

Review time did not differ significantly in the first and the second round for both charts (mean [SD] = 72.8 minutes [45.6] in first round and 61.0 minutes [47.6] in second round) and profiles (32.3 minutes [14.0] and 28.3 minutes [16.3] respectively).

## DISCUSSION

In this study, we examined application of the clinical reasoning process to structured patient data for phenotype evaluation. It has long been posited that crucial information about the patient state, diagnoses and symptoms is most fully and accurately recorded in unstructured free-text notes and that only the notes can serve as the gold standard in phenotype evaluation. Indeed, unstructured data offers great opportunity for expression, allowing clinicians to both interpret other providers’ narratives and create their own [31]. As a result, there have been multiple disease-specific endeavors in natural language processing aiming at improving phenotype development and evaluation by capturing richness of free text [27,46–48].

KEEPER mimics interpretation of free-text narratives about patient state and can complement probabilistic methods for phenotype evaluation [30] by providing scalable yet transparent and interpretable solution for status ascertainment. As we demonstrate here, standardizing data representation according to the elements of clinical reasoning enables effective sense-making. Using structured data alone, clinicians can construct narratives that align well with explicitly written narratives in charts. The efficiency of this process depends on the ability of structured data to reflect true patient state and its ability to reduce cognitive load.

KEEPER is efficient if the structured data contain the necessary elements for valid inference and therefore its performance may depend on comprehensiveness of data capture and specifics of patient population in a given data source. Data is likely to be sufficient to infer prevalent conditions and conditions requiring drug therapy or operative procedures [49–51]. On contrary, it is commonly acknowledged that asymptomatic conditions and some co-morbidities are underrepresented in structured data [52]. Similarly, structured data and billing codes are not likely to capture conditions associated with privacy concerns [53]. It is not clear to what extent the performance observed in this study can be replicated on claims data sources for those conditions whose diagnosis is heavily measurement-based. In our example, sensitivity of KEEPER may be low when attempting to classify patients with COPD or ESRD on claims data sources as there are patients who do not receive specific treatment and, therefore, can be misclassified as controls.

As the goal is not a comprehensive patient evaluation but rather case adjudication in respect to one specific disorder, presenting only relevant information is required to efficiently process the information about the patient. Patient structured record can contain hundreds and thousands of events, codes, and values, which decreases efficiency of review and increases the likelihood of missing important information. KEEPER represents only relevant data in a structured way, which decreases time to review and improves accuracy and agreement between the reviewers and supports previous findings on benefits of standardized practices for patient ascertainment [54,55].

On the contrary, high volume of information and contradicting information in charts were a source of disagreement among reviewers. For example, COPD has to be differentiated with asthma, which requires assessment of history of disease, pulmonary tests and previous drug exposures.

In our patient sample, some patients with bronchial obstruction did not have history of asthma in the recent notes but previous notes (sometimes going back 10 years and more) had a diagnosis of asthma, montelukast (a drug almost exclusively used for mild and intermittent asthma) or bronchodilator use, which undermined the reliability of the later diagnosis of COPD. Finding this information required scrutinizing tens of clinical notes, which lengthened reviews and decreased accuracy.

We proposed that standardizing the input and output of KEEPER facilitates scalability of chart review as the former has a potential to perform similarly across a broad range of conditions. While examining this hypothesis on all possible conditions is not feasible, we selected a mix of chronic and acute, inpatient and outpatient conditions to cover a variety of conditions. On one hand, we observed consistent improvement in inter-rater reliability across all conditions, which strengthened our assumption that KEEPER can be seen as a disease-agnostic solution. On the other hand, variable accuracy of review across different conditions points at the need for more research on factors influencing inference from structured data. In COPD, the factors that contributed to lower accuracy compared to other conditions included inability to (a) easily interpret the results of pulmonary function tests to distinguish COPD from asthma or chronic bronchitis and (b) ascertain the cases when no results of pulmonary function tests were available. Similar challenges were encountered in full chart review, especially if the results of pulmonary functions tests were contradictory or inconclusive.

Disagreement among reviewers in patient ascertainment using full charts can partially be explained by differences in clinical training and expertise and different approaches to chart review. One scenario used by clinicians involved starting with the day 0 provided to clinicians and reviewing patient data around the day 0 first, moving sequentially along the longitudinal patient record. Another scenario involved starting at the data elements that carried the most accurate perceived information (such as pathology reports for acute appendicitis or specialty notes associated with laboratory values for the other conditions) and then retrospectively reconstructing the case. Standardization of data representation in KEEPER partially mitigated this issue leading to higher inter-rater reliability.

As we noted, reviewing profiles was substantially faster and the time spent on profile review was relatively consistent across the cases and rounds. Therefore, researchers can review more patients with KEEPER, thus enabling more reliable estimation in clinical studies. It can be especially useful in patient adjudication for safety research where rare outcomes require larger sample sizes [56].

In the future, we envision KEEPER as a user interface integrated in a broader stack of OHDSI tools, which will enable seamless integration of phenotype development, cohort execution, cohort diagnostic and phenotype evaluation [30,57–59]. For this solution to be scalable, relevant information must be extracted in an automated disease-agnostic fashion. There are many works on identifying similar concepts, including lexical, ontological and data-driven approaches [60–62] to can be leveraged to accomplish this task. Given complexity of the task, an appropriate method should be able to identify relevant but not necessarily semantically similar concept, concepts from different domains (such as laboratory tests relevant to a given disease) and clinically meaningful concept pairs (such as diagnosis-differential diagnosis pairs [63]).

## LIMITATIONS

Our findings may not be generalizable to the institutions with higher expected information loss from charts to structured records. We conducted the experiment for four conditions and while these conditions represent a spectrum of disorders requiring different levels and settings of care, the results may not be generalizable to other conditions. We also enlisted only four clinicians and clinician performance and experience may vary.

## CONCLUSIONS

Phenotype evaluation remains the bottleneck of observational research the current gold standard - chart review - is interpretable and generally trusted but expensive, time-consuming, and variable. In this study, we evaluated the ability of structured data to support effective patient status ascertainment. We used the principles of clinical reasoning, standardization, and dimensionality reduction to build a knowledge-enhanced patient profile review system or KEEPER. We demonstrated that structured data can support valid inference about patient state if organized and presented according to these principles. KEEPER showed similar accuracy and higher inter-rater reliability compared to chart review at a fraction of time.

## Data Availability

The data used in this study are protected health information and are not available

## IRB statement

We obtained an approval to conduct this research from the Columbia University Medical Center institutional review board (IRB-AAAS6414).

## Authors’ contribution

AO, GH and PR contributed to conception and design of the study and its interpretation. AO, GH, SAH, AE, LR and MS participated in the experiment and contributed to experimental data acquisition. AO executed the study and performed data analysis. All authors participated in manuscript writing, read and approved the final manuscript.

