## Supplementary Table 1 for "Development and evaluation of a scalable alternative to chart review for phenotype case adjudication using standardized structured data from electronic health records"

**Supplementary Table 1.** Examples of conceptual elements for selected acute and chronic, inpatient and outpatient conditions.

| **Condition and its characteristics** | **Demographics and details about the visit** | **Presentation** | **Prior conditions, symptoms, and treatment** | **Diagnostic procedures** | **Laboratory tests** | **Competing diagnoses** | **Treatment procedures and medications** | **Complications** |
| --- | --- | --- | --- | --- | --- | --- | --- | --- |
| Acute appendicitis;  Acute, treated in inpatient settings | More common in young patients, slightly more females | Right lower quadrant pain, loss of appetite, nausea and vomiting, stomach distress, fever | None, acute onset; family history of appendicitis | Ultrasound of abdomen, computer tomography of abdomen, x-ray of abdomen | Leukocytosis | Diverticulitis, ileitis, Crohn's disease , ovarian abscess or cyst, endometriosis in women | Appendectomy (immediate or interval) or intravenous broad-spectrum antibiotics followed by oral | Perforation, sepsis |
| Chronic obstructive pulmonary disease;  Chronic, treated in inpatient and outpatient settings | More common in men and adults older than 40 | Cough, wheezing, shortness of breath, decreased exercise tolerance, tachycardia | Smoking, dust exposure, bronchitis, emphysema, heart failure | Pulmonary function tests (spirometry), chest x-ray, Pulse oximetry and arterial blood gases | Respiratory acidosis | Asthma, lung cancer, interstitial lung disease, bronchiectasis, tuberculosis | Short- and long-acting beta agonists, long-acting muscarinic antagonist, inhaled and oral glucocorticosteroids | Pulmonary hypertension, right heart failure, |
| Chronic, treated in inpatient and outpatient settings | Kids and adolescents, both genders | Polydipsia, polyuria, weight loss, hyperglycemia, ketonemia, ketonuria, perineal candidiasis | Exposure to Coxsackie B virus, family history of type I diabetes |  | Elevated plasma glucose, both random and fasting, elevated glycated hemoglobin, presence of pancreatic autoantibodies, low fasting insulin, low C-peptide | Type II diabetes mellitus, pancreatic diabetes, hyperglycemia in other conditions | Various types of insulin | Neuropathy, nephropathy, cataracts, retinopathy, ketoacidotic and hypoglycemic coma |
| Chronic, treated in inpatient and outpatient settings | Both genders, more likely in young men/elderly women | Seizure | May be prior neurological damage: stroke, traumatic brain injury, infectious disease of brain | Electroencephalography, neuroimaging, lumbar punction |  | Febrile seizures, eclampsia, brain tumor, acute stroke or hemorrhage | Long course of non-gabapentinoid anti-seizure medications (felbamate, levetiracetam, lamotrigine, clobazam, etc.) and gabapentinoid medications (Pregabalin and gabapentin) | Increased risk for personal injury, accidental death, and drowning as well as psychiatric comorbidity, suicidal deaths, and sudden unexpected death in epilepsy. |
| Chronic, treated in inpatient and outpatient settings | Both genders | Nausea and vomiting, fatigue, hypertension, shortness of breath, edema | Chronic kidney disease, glomerulonephritis, polycystic kidney disease, diabetes, hypertensive disorder | Ultrasound of kidneys, computer tomography of kidneys | Decreased estimated glomerular filtration rate | Acute kidney failure, other stages of chronic kidney disease | Peritoneal and hemodialysis, kidney transplantation, symptomatic care | Anemia, hyperkalemia, osteoporosis, immunodeficiency, pericarditis, uremic pneumonitis |
| Chronic autoimmune hypothyroidism;  Chronic, treated in outpatient settings | More common in women | From asymptomatic to fatigure, weight gain, myalgia, constipation, goiter, bradycardia, hypotension | Hypertyroidism with subsequent treatment with high doses of methimazole or propylthiouracil | Ultrasound of thyroid gland, magnetic resonance imaging of brain | Elevated serum thyroid-stimulating hormone and low or normal serum free thyroxine, elevated thyroid peroxidase antibodies | Pituitary adenoma, external neck irradiation (Hodgkin lymphoma, neck cancer), drug-induced hypothyroidism | Liothyronine, levothyroxine | Myxedema coma |
| Acute bacterial sinusitis;  Acute, treated in outpatient settings | None | Rhinitis, facial pain, headache, hyposmia or anosmia, fever | None; risk factors include immunodeficiencies, smoking, dental infections | Computed tomography or magnetic resonance imaging of head in complicated cases | Microbiology culture of sinus aspirate | Viral sinusitis, common cold, other types of headaches, dental conditions | Analgesics and antipyretics, broad-spectrum oral antibiotics | periorbital and orbital cellulitis, osteomyelitis of sinus bones, meningitis |
| Acute myocardial infarction;  Acute, treated in inpatient settings | More common in older adults | Chest pain or irradiating pain, shortness of breath, | Chronic stable angina, hyperlipidemia, metabolic syndrome, smoking | Electrocardiogram, angiography, | Elevated troponin | Pulmonary embolism, aortic dissection, pericarditis, cardiomyopathy, gastrointestinal disorders (acute pancreatitis, ulcer disease, etc.), disorders of chest wall and spine | Percutaneous coronary intervention, fibrinolysis, anticoagulants, beta-blockers, nitrates | Cardiogenic shock, heart failure, arrythmias, ventricular aneurysm |
| Acute pancreatitis;  Acute, treated in inpatient and outpatient settings | None | Epigastric pain, nausea and vomiting | Alcohol dependence, cholangitis | Endoscopic ultrasonography, ultrasound examination of abdomen, computer tomography of abdomen | Elevated lipase and amylase | Gastric or duodenal ulcer, intestinal obstruction | Fluid replacement, pain control, nutrition management, cholecystectomy | Pancreonecrosis, steatorrhea, diabetes, pancreatic calcification, and fibrosis |
| Prostate cancer;  Chronic, treated in inpatient and outpatient settings | Males, median age is 65 | Varies from asymptomatic (diagnosed during screening) to urinary tract obstruction, nocturia, incomplete bladder emptying to pathologic fracture and symptoms of metastatic disease | prior symptoms of urinary obstruction | Transrectal ultrasound, prostate MRI, prostate biopsy | Elevated prostate-specific antigen | adenoma of prostate, urinary tract infection, bladder cancer, prostatitis | Radical prostatectomy, neoadjuvant androgen deprivation therapy, radiation therapy in combinations, active surveillance for some groups | Metastatic neoplastic disease |
